# Analysis and Augmentation of Small Datasets with Unsupervised Machine Learning

**DOI:** 10.1101/2021.04.21.21254796

**Authors:** Serge Dolgikh

**Affiliations:** Dept. of Information Technology, National Aviation University

**Keywords:** Unsupervised Learning, Clustering, Ensemble learning, statistical analysis, small data

## Abstract

Analysis of small datasets presents a number of essential challenges not in the least due to insufficient sampling of characteristic patterns in the data making confident conclusions about the unknown distribution elusive and resulting in lower statistical confidence and higher error. In this work, a novel approach to augmentation of small datasets is proposed based on an ensemble of neural network models of unsupervised generative self-learning. Applying generative learning with an ensemble of individual models allowed to identify stable clusters of data points in the latent representations of the observable data. Several techniques of augmentation based on identified latent cluster structure were applied to produce new data points and enhance the dataset. The proposed method can be used with small and extremely small datasets to identify characteristics patterns, augment data and in some cases, improve accuracy of classification in the scenarios with strong deficit of labels.

## 1 Introduction

Analysis of small datasets presents a number of essential challenges not in the least due to insufficient sampling of characteristic patterns in the data making confident conclusions about the unknown distribution elusive and resulting in lower statistical confidence and higher error. On the other hand, it can be essential in the cases and scenarios of novel and / or rare conditions where large amounts of data do not exist, or have not been accumulated [1].

One of the known challenges in application of machine learning methods, including artificial neural networks to small datasets is that of the stability of learning. It can manifest itself in strong dependency on the choice of parameters, selection and order of training batches and other factors [2,3], as well as overfitting and inability to generalize. Due to this small data volatility results obtained with models of similar architecture with the data can be inconsistent, and the performance in generalization and accuracy, limited. As well, reproducibility of the results can be problematic even with the same architecture and data, complicating comparison, improvements and optimization.

Earlier studies attempted to approach the problem of stability of machine learning with small data with a variety of methods such as k-fold cross-validation and ensemble methods [4]; Radial-Basis Function (RBF) neural networks [5,6] and a number of other methods [7,8]. However, while many of these methods were successful in specific applications, generality in application to different data and problems to the best of our knowledge could not be assured due, not in the least, specifics of the architecture and / or essential assumptions about distribution of the data.

On the other hand, methods of unsupervised machine learning [9,10] have shown an effective ability to achieve significant reduction of dimensionality, or redundancy of the observable parameter space that in a number of cases were instrumental in the analysis and determination of characteristic patterns and trends in complex data [11-13] including constrained data [14]. Importantly, application of these methods does not require data labeled with confidently known outcome and generally can be performed with smaller samples of data. In our view, these characteristics make these methods good candidates for an analysis of early and rare conditions, scenarios and situations, where large amounts of confident data have not yet been accumulated, while still allowing to aggregate data for application of conventional methods of statistical analysis at later stages.

To address the outlined challenges in the analysis of small datasets, it was proposed to use an ensemble of unsupervised models to identify characteristic structure in the input data, addressing both problems of the label deficit and stability of training with minimal datasets. The structure in the latent representations of unsupervised generative models that can be identified with this approach can then be used to produce new data points, generated based on identified characteristics of distribution of data points in the latent representation. Unlike some of the discussed approaches, this method does not depend on specific assumptions about the distribution of the data and can be used with data of any origin.

## 2 Methodology

An ensemble of unsupervised artificial neural networks with the architecture of deep autoencoder with strong dimensionality reduction [10] was used to produce low-dimensional (namely, two or three-dimensional) representations of a small dataset of real diagnostics data, as described further in this section. The data was obtained from openly available statistics of the early phase Covid-19 epidemics, by national and subnational jurisdiction. The advantages of the selected architecture are: demonstrated efficiency in producing informative low-dimensional representations of diverse real-world data as well as universal approximation capacity of neural networks [15] making them suitable for virtually any type of data.

### 2.1 Unsupervised Autoencoder Neural Network

A deep autoencoder neural network model had several deep layers and a central encoding layer of size two, producing a two-dimensional latent representation defined by axes representing activations of neurons in the encoding layer. The decoding / generating stage was fully symmetrical to the encoder. An architecture diagram of the model is shown in Figure 1.

**Fig. 1.**
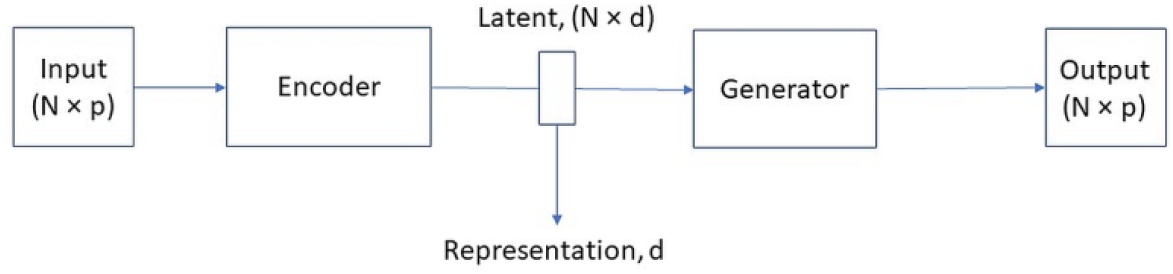
Deep autoencoder with dimensionality reduction (p = 9, d = 2).

Overall, the model had five fully-connected deep layers and approximately 8,000 trainable parameters. For detailed description of the model architecture refer to [14].

Unsupervised training was performed in an unsupervised process with minimization of deviation of the output of the model from the input with binary cross-entropy cost function. Training over 30 – 50 epochs produced noticeable reduction in the value of the cost function for majority of the training models, indicating the success of unsupervised training.

### 2.2 Data

A small dataset of approximately 40 cases of early Covid-19 national epidemiological statistics (Early Covid-19 Epidemiology, ECE) was used, compiled from sociological and epidemiological statistic obtained from open sources [14]. The data described observable factors characterizing the state of selected national and subnational jurisdictions at the time of early exposure to Covid-19 pandemics with nine observable parameters, including: population and demographics factors, epidemiological practice, public health policy and others, as described in detail in [14], Table 1.

**Table 1.** Description, ECE dataset

| Parameter | Size | Dimension-ality | Range | Mean, EI<sup>1</sup> | Time scale |
| --- | --- | --- | --- | --- | --- |
| Value | 41 | 8 | 0.0 – 1.0 | 0.36 | 3 months |

<sup>1</sup> Epidemiological impact.

### 2.3 Ensemble of Unsupervised Models

An ensemble approach with a set of pre-trained models of the architecture described in the previous sections was used to improve the confidence of the analysis, given the small size of data and the outlined challenges. A group of thirty models of the architecture described in Section 2.1. was pre-trained with the dataset in an unsupervised process based on minimization of generative error. Trained models produced two-dimensional representations of the dataset illustrated in Figure 2.

**Fig. 2.**
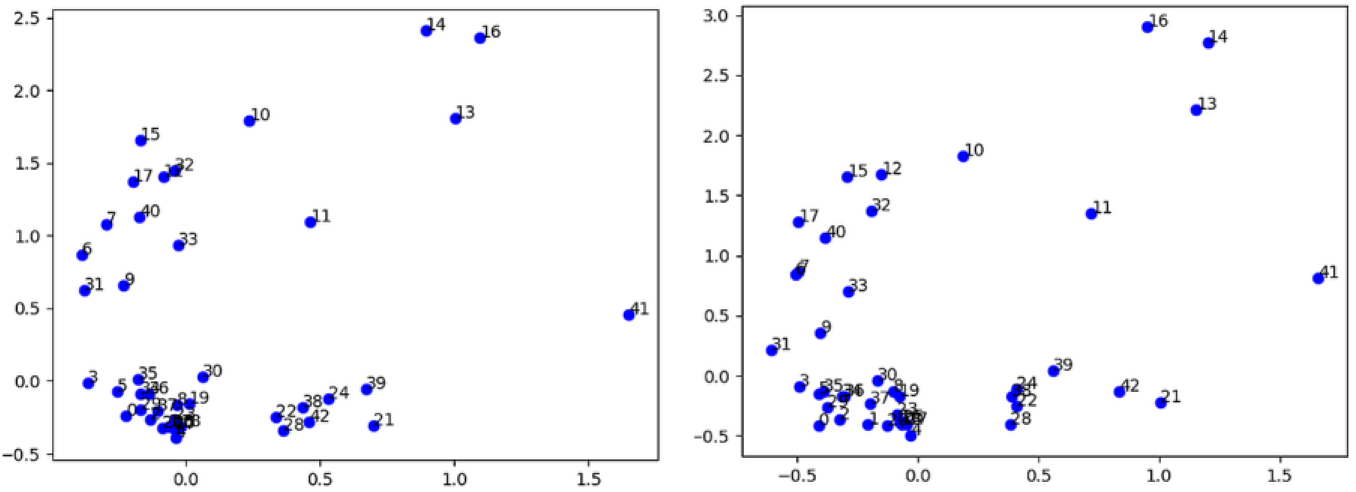
Latent distributions of ECE data with deep autoencoder models.

From the resulting ensemble of pairs (trained model, latent distribution) a subset with higher effectiveness of generative learning was selected by the following criteria:

- Training accuracy, measured by the change and final value of the cost function;
- Clusterization performance, evaluated visually from distribution of data in the latent axes (example in Fig.2).

By the first criterium, only the models with final cost below certain threshold were retained. The latter was identified visually in a blind process, that is, the contents of clusters in the latent distribution were not examined, only the capacity of the model to produce clearly identifiable regions with higher concentration of data points (Fig.2). In the future research, this phase of the selection process can be automated via application of methods of unsupervised clustering such as DbScan [16] and similar.

The resulting subset of five models was used to analyze the distributions of data in the latent representations and evaluation of the method of unsupervised augmentation of small datasets with generation of new data points based on identified latent cluster structure.

## 3 Results

### 3.1 Consistency of Cluster Content

An analysis of distributions in the selected ensemble of effective generative learning as described in the previous section produced an essential finding: though latent distributions showed significant variance between individual models, the contents of the clusters identified in the latent representations were mostly stable between models in the ensemble.

As individual models were selected blindly, based on clusterization quality but not the content of specific clusters, a priori there could be no expectation that clusters produced by different models should have the same or similar composition. It is more likely though in the case where identified clusters represent stable patterns in the observable data and the models were successful in learning them under the incentive to produce good quality generation of the original data. Table 2 shows characteristics of latent clusters identified with the unsupervised ensemble method.

**Table 2.** Latent clusters, ECE dataset

| Cluster | Population | Variation,<br>population | Latent area * | Variance,<br>latent area |
| --- | --- | --- | --- | --- |
| Cluster 1 | 9 | ~ 22 % | 0.16 | 0.04 |
| Cluster 2 | 21 | ~ 15% | 0.21 | 0.05 |

\* Relative to the total latent area of the dataset

Stability of latent clusters is a key foundation of the method of unsupervised augmentation of small data, indicating that clusters described significant patterns in the dataset.

### 3.2 Cluster-based Data Generation

The architecture autoencoder allows a direct method of propagating a position in the latent representation to the observable (i.e. input) parameters. It can be achieved by using a latent position with coordinates *L = (l*_*1*_, *l*_*2*_) as an input to the generating part of the neural network model (Fig.1) to obtain the output *X*_obs_ in the observable parameters:

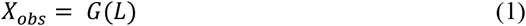

where *G: R → O*, is generating submodel of the autoencoder operating from the latent representation *R* to the observable space *O*.

Generative capacity of unsupervised models thus allows to generate new data points by selecting specific latent positions in the representation space and obtaining their observable images in the coordinates of the input parameters with (1).

Based on the observation in Section 3.1 on the stable content of the identified clusters, the described process can be applied to obtain new data points with observable characteristics similar to those in the identified clusters as follows:

- In the first step, generating model(s), one or several, are selected based on the criteria of learning quality as described previously;
- In the latent representations of selected models, areas corresponding to identified latent clusters are defined, based on stability of cluster content as discussed in Section 3.1.
- In the latent regions associated with identified clusters, new latent positions are selected, by a number of methods.
- The selected positions are propagated to the observable space with (1), producing a new set of data points with the coordinates corresponding to observable parameters.
- In the final step, the generated data points can be added to the original dataset, resulting in non-trivial augmentation.

The effectiveness of the method of unsupervised clustering augmentation can be supported by the following considerations:

Suppose there is a small dataset *S* of size *N* with *P* observable parameters. If a conventional approximation method was used, such as Gaussian, the error of the mean in each of the parameters could be estimated as mean *p / √N*. With a low-dimensional latent representation, in the case of good clusterization, the original distribution can be approximated by a quasi-multi-modal distribution with the number of modes *d × Nc, d* being the latent dimensionality, where *Nc* the number of clusters, with considerably smaller error of mean and standard deviation, as long as *d* remains sufficiently small and the number of samples in the principal clusters, sufficiently large.

Figure 3 demonstrates application of the augmentation method with ECE dataset.

**Fig. 3.**
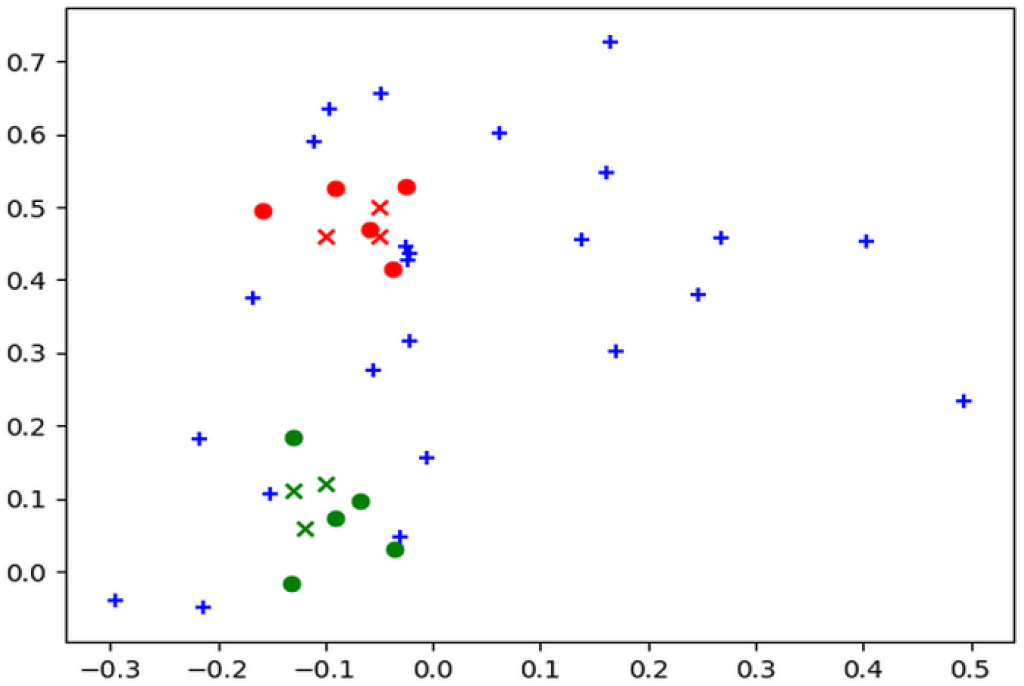
Cluster-based data augmentation. Green, red: identified latent clusters, cross: new latent data points.

### 3.3 Model Invariance of Generated Data

An important question in application of the proposed small data augmentation method is how stable the generated data is with respect to the latent position of origin, given that latent representations are specific to individual models?

Usability of the method thus depends on the assumption that generated observable data would be mostly consistent between individual generating models, that is, in some way, model-invariant. Given the essential nature of this assumption, it was verified by using the augmentation method with two different model instances.

It was observed that while latent representations produced by models were essentially different by positioning of data points in the latent area, generation of new data points from positions in the neighborhood of real data points, that can be considered as “anchor points” of the latent – observable transformation resulted in similar observable output. In Table 2, the Euclidean distance measured in the observable coordinates between data points generated from the latent neighborhood of a real data point in the identified clusters (anchor point) with two different generating models is compared with characteristic distances of data distributions in the clusters and the entire dataset.

**Table 2.** Generated data – model invariance

|  | Max distance,<br>generated | Max distance, cluster | Max distance, dataset |
| --- | --- | --- | --- |
| Cluster 1 | 0.05 | 0.825 | 1.0 |
| Cluster 2 | 0.02 | 0.914 | - |

These results demonstrate that the method is to a large extent invariant with respect to selection of the generating model from the ensemble and can be used reliably to generate new data with characteristics determined by and similar to significant patterns in the original dataset. Note that the results above were produced with multivariate cubic interpolation [17] from identified stable data points in the clusters and their latent positions rather than generative model (1) due to relative simplicity of interpolation with a small number of observable parameters providing similar or better accuracy whereas optimization of the neural network architecture for generative accuracy was not the main focus of the study.

## 4 Applications

### 4.1 Augmentation of Small Data

To summarize the results of the previous sections, the method of augmentation of small dataset with an ensemble of generative models can be applied under the following conditions:

- The latent dimensionality is sufficiently small.
- The models show good learning accuracy and clear and consistent latent clusterization.
- The number of identified latent clusters is small compared to the number of samples in the dataset, and the population of main clusters is sufficiently large.

Under these conditions, the original data can be reliably described and augmented with a multi-modal distribution in the latent regions of identified clusters as exemplified in the results of the study.

### 4.2 Classification

The method of augmentation of small datasets can be used to improve results of classification of supervised learning models trained with small dataset. In regular practice, the size and representativity of the training data can have strong influence on the resulting efficacy of classification.

The application is not unconditional though and essentially depends on existence of a correlation of principal clusters and the effect of interest, that can be used as a label in supervised learning and classification. If such a correlation can be observed in the original data, augmentation of the dataset with new data points with labels assigned based on cluster relation can produce significant improvement in classification accuracy. In the dataset used in the study, clusters appeared to have a correlation with the factor of interest (i.e., epidemiological impact per jurisdiction), with identified clusters 1 and 2 associated, by preliminary results, with higher-impact and average scenarios respectively, though detailed investigation of correlation was not the primary objective of this study and will be examined in more detail elsewhere.

However, if correlation of cluster to factor of interest cannot be established reliably, it is possible that the latent coordinates in which clusterization was observed may not have direct correlation to the effect of interest, and a different method of labeling should be used, though increasing the size of the dataset may still produce improvements in classification accuracy.

## 5 Discussion

The approach based on an ensemble of unsupervised generative models to determine characteristic clusters with a small dataset of real diagnostics data demonstrated in this work has proven its effectiveness in the in the considered case. The clusters identified with unsupervised methods allowed to generate new data points with high level of confidence, with a number of possible applications, including augmentation of general datasets and in some cases, as discussed in Section 4.2, to improve classification performance.

In novel and / or rare cases accumulation of volumes of data needed for confident analysis with conventional methods of machine learning could present a serious challenge. Application of methods of unsupervised machine learning, with identification of characteristic patterns even with smaller datasets can offer a direction toward improving the confidence and accuracy in the analysis of small data.

## Data Availability

Data used in the study is available upon request

https://www.medrxiv.org/content/10.1101/2020.05.17.20104661v2

https://www.google.com/covid19-map/

https://www.worldometers.info/

## References

1. Hekler E.B., Klasnja, P., Chevance G. et al.: Why we need a small data paradigm, BMC Med., 17 133 (2019).

2. Wasserman P.D.: Neural computing: theory and practice. Van Nostrand-Reinhold, New York (1989).

3. LeBaron B., Weigend A.S.: A bootstrap evaluation of the effect of data splitting on financial time series. IEEE Trans. Neural Networks 9 213–220 (1998).

4. Cunningham P., J. Carney, S. Jacob S.: Stability problems with artificial neural networks and the ensemble solution. Artificial Intelligence in Medicine, 20 (3) 217–255 (2000).

5. Karar M.E., Robust RBF neural network-based backstepping controller for implantable cardiac pacemakers, Int. J. Adap. Cont. Sign. Proc 32 1040–1051 (2018).

6. Izonin, I., Tkachenko R., Dronuyk I. et al.: Predictive modeling based on small data in clinical medicine: RBF-based additive input-doubling method. Math Biosc. Eng, 18 (3) 2599–2613 (2021).

7. Forman G., Cohen I.: Learning from little: comparison of classifiers given little training. In: Proceedings of PKDD, 19 161–172 (2004).

8. Geris L.: Computational modeling in tissue engineering. Springer-Verlag, Berlin (2013).

9. Fischer, A., Igel, C.: Training restricted Boltzmann machines: an introduction. Pattern Recognition 47, 25–39 (2014).

10. Bengio, Y.: Learning deep architectures for AI. Foundations and Trends in Machine Learning 2(1), 1–127 (2009).

11. Coates, A., Lee, H., Ng, A.Y.: An analysis of single-layer networks in unsupervised feature learning. In: Proceedings of 14th International Conference on Artificial Intelligence and Statistics 15, 215–223 (2011).

12. Rodriguez, R.C., Alaniz, S., and Akata, Z.: Modeling conceptual understanding in image reference games. In: Advances in Neural Information Processing Systems (Vancouver), 13155–13165 (2019).

13. Prystavka, P., Cholyshkina, O., Dolgikh, S., Karpenko, D.: Automated object recognition system based on convolutional autoencoder. In: 10th International Conference on Advanced Computer Information Technologies (ACIT-2020), Deggendorf, Germany, 830– 833 (2020).

14. Dolgikh, S.: Identifying explosive epidemiological cases with unsupervised machine learning. In: Proc. 3rd International Conference on Informatics & Data-Driven Medicine, Vaxjo Sweden (2020).

15. Hornik K., Stinchcombe M., White H.: Multilayer feedforward neural networks are universal approximators. Neural Networks 2(5), 359–366 (1989).

16. Ester, M., Kriegel, H-P., Sander, J., et al.: A density-based algorithm for discovering clusters in large spatial databases with noise. Proc. Second International Conference on Knowledge Discovery and Data Mining (KDD-96) 226–231 (1996).

17. Wendland, H.: Scattered data approximation. Cambridge University Press 9 (2005).

